# A Mobile AI-enhanced Platform for Standardized Wound Assessment and Clinical Decision Support

**DOI:** 10.64898/2026.01.22.26344407

**Authors:** Mahla Abdolahnejad, Nikoo Mashayekhi, Manuella Kyremeh, James Smith, Moniz Chan, Gloria Fang, Thinoj Jegatheeswaran, Hannah O. Chan, Rakesh Joshi, Colin Hong

## Abstract

Chronic wounds affect over 1.2 million Canadians and incur healthcare costs exceeding $13 billion annually, with global expenditures approaching $149 billion. Current clinical practice relies on manual measurements and subjective visual evaluations, which overestimate wound area by up to 40% and demonstrate poor-to-moderate inter-rater reliability. This variability complicates longitudinal monitoring and evidence-based treatment selection. We developed and evaluated an integrated mobile platform combining deep learning-based wound assessment with clinical decision support. A curated dataset of 1,648 de-identified clinical wound photographs was assembled from wound care clinics, representing diverse aetiologies (arterial, venous, diabetic foot ulcers, pressure injuries) and skin tones (32% Monk Skin Tone 7-10). Three convolutional neural networks were trained: (1) an EfficientNet-B7-based classifier for wound etiology, (2) a gated pressure injury staging network, and (3) a DeepLabv3 encoder-decoder architecture with ResNet backbone for multi-class tissue segmentation (epithelialization, granulation, slough, eschar). Fiducial marker-based calibration enabled automated wound size quantification. A rule-based recommendation engine mapped assessment outputs to evidence-based dressing selections. The system was deployed as a cross-platform mobile application with cloud-native backend infrastructure. The wound classification model achieved 91.75% mean accuracy across four wound categories. Pressure injury staging accuracy ranged from 67% (Stage III) to 92% (Stage I). Tissue segmentation yielded a mean Dice similarity coefficient of 0.64 ± 0.06 and pixel-level accuracy of 98%.

Automated size estimation demonstrated strong correlation with manual measurements (r = 0.73, n=53), with mean absolute error of 3.7 ± 2.1 mm; 84.2% of measurements fell within the ±5 mm clinical equivalence margin. Fiducial marker detection succeeded in 93% of test images. Performance remained stable across skin tone categories and imaging conditions. This integrated platform demonstrates technical feasibility for standardized, objective wound assessment addressing documented limitations of manual practices. The system provides interpretable segmentation overlays and actionable treatment recommendations while maintaining clinician oversight. These findings support progression to prospective validation studies evaluating real-world clinical utility and patient outcomes.

## INTRODUCTION

Pathological wounds in North America are described as a hidden epidemic with significant socioeconomic implications. Recent data indicates that in Canada alone, more than 1.2 million individuals (3 % of population) are affected by acute and chronic wounds, leading to wound care expenses surpassing $13 billion in 2024, with a disproportionate effect on rural communities (Green & Botros, 2025; Freeman et al., 2025). Globally, the wound care expenditures reached approximately $149 billion in 2022, with significant costs attributed to managing chronic wounds (Sen, 2025). These types of wounds, such as diabetic foot ulcers, venous and arterial ulcers, pressure injuries, and non-healing surgical wounds, are identified by long healing processes and uncertain outcomes (Wilkinson & Hardman, 2020; Wu et al., 2022). Older adults are disproportionately impacted due to age-related vascular issues, reduced skin elasticity, and higher rates of chronic conditions like diabetes (McDermott et al., 2022).

Chronic wounds (also termed difficult-to-heal wounds) not only incur significant healthcare expenses but are also often overlooked in data analysis, frequently misclassified as secondary effects of other illnesses rather than being recognized as distinct conditions requiring specialized attention and research investment, contributing to limited funding for wound care and lack of education regarding wound physiology and treatment (Sen, 2009). For individuals, these types of wounds greatly diminish quality of life by limiting mobility, causing persistent pain, and increasing the risk of infections. For diabetic foot ulcers specifically, the morbidity rate post-ulceration is notably high, with recurrence rates of 65% within 3-5 years after healing and a lifetime risk of lower-extremity amputation close to 20% (McDermott et al., 2022).

Accurate and consistent assessment of wounds is crucial for guiding treatment decisions, tracking healing progress, and reducing complications, including the use of photographs (Loizou et al., 2012). Key clinical factors include wound size, tissue composition (granulation, epithelialization, slough, necrosis), infection status, and treatment response. Currently, clinical practice heavily relies on manual measurements and subjective visual evaluations, leading to significant variability and measurement inaccuracies (Martinengo et al., 2019). Manual measurement methods often overestimate wound area by up to 40% and show poor to moderate reliability between different assessors (Jorgensen et al., 2016Khoo & Jansen, 2016). Even trained nurse-assessors demonstrate significant disagreement when assessing wound infection, emphasizing the need for standardized, objective evaluation tools. This inconsistency in wound assessment complicates long-term monitoring, hampers clinical decision-making, and obstructs evidence-based treatment selection.

To address these challenges, digital health platforms and tele-wound care models have been developed to improve standardization of wound assessment, enhance specialist consultations, and streamline post-care (Mohammed et al., 2025). However, systematic assessments reveal that available patient-oriented mobile wound care apps remain limited in quality and adequate apps for patients with chronic wounds are scarce (Dege et al., 2024). Digital planimetry systems offer automated wound measurements with better precision than manual methods through techniques such as image preprocessing, Otsu’s thresholding for segmentation, and DPI-based scaling for clinical measurements (Ferreira et al., 2021), although they are limited to two-dimensional analyses and may not capture crucial parameters like wound depth (Khoo & Jansen, 2016).

Advanced computational techniques, particularly machine learning (ML) and deep learning models using convolutional neural networks (CNNs), show promise in automating comprehensive wound assessments. Recent reviews compile evidence indicating that AI-driven systems can accurately outline wound boundaries, classify tissue types, and estimate key parameters, though current smartphone applications for wound assessment face major limitations including lack of algorithm transparency and insufficient clinical validation data, which hinder clinical adoption (Griffa et al., 2024). Comprehensive reviews of mobile apps for wound assessment reveal significant limitations alongside emerging advancements and opportunities in this rapidly evolving field (Kabir et al., 2024). Current status assessments of AI applications in wound repair indicate both considerable challenges and promising prospects for the field (Liu et al., 2025). Texture-based image analysis is also emerging as a reliable approach for objective wound monitoring, with studies demonstrating that quantitative texture features can act as dependable markers for tracking healing progress (Loizou et al., 2012).

Several recent studies focus on segmenting wound tissues into meaningful classes, such as epithelialization, granulation, slough, and eschar. Models trained on specific clinical datasets can consistently quantify tissue proportions at the pixel level, reducing observer bias and enhancing measurement consistency (Ramachandram et al., 2022). Research into inter-rater agreement highlights the inherent inconsistency in manual annotations, emphasizing the need for algorithmic standardization and robust protocols for establishing reference datasets (Morgado et al., 2025). Moreover, studies on device validation and workflow integration demonstrate that AI-assisted, smartphone-based solutions exhibit reliability across various platforms and imaging settings, indicating their feasibility for real-world clinical application (Mohammed et al., 2025). Complementary technologies, like bacterial fluorescence imaging devices, have proven useful in achieving high accuracy (>95%) in wound measurements and providing guidance for targeted debridement, showcasing the broader potential of device-supported wound care (Raizman et al., 2019).

A critical factor for the clinical implementation of AI algorithms in wound care is their interpretability and transparency. Post-analysis attention mechanisms and saliency methods, such as Class Activation Mapping (CAM) and Gradient-weighted CAM (Grad-CAM), are increasingly used to visualize image regions that influence model predictions. However, systematic meta-analyses reveal persistent gaps in user-friendly evaluation, methodological transparency, and ethical considerations, with notable absence of research evaluating explainability, clinician trust, or usability in real-world settings, emphasizing the necessity of longitudinal clinical validation, participatory system design, and uniform interpretability measures for responsible AI implementation in healthcare (Abbas et al., 2025). In the realm of medical imaging and particularly wound-specific applications, these explainability methods enhance trust and usability when appropriately integrated (Brima & Atemkeng, 2024). However, their reliability and quantitative rigor require thorough validation. Hence, it is crucial to combine robust baseline performance (accuracy in segmentation and classification) with transparent visual explanations and well-documented failure modes.

This study expands on existing advancements made by the research group on AI-enabled medical assessment applications for Burns and Vitiligo (Lee et al., 2025, Abdolahnejad et al., 2025 for Burns; Abdolahnejad et al., 2024 for Vitiligo). As described, the study introduces a comprehensive wound assessment system that integrates various components, including image standardization, CNN-based inference for wound classification, semantic segmentation of wound tissues, and a decision layer for generating wound dressing recommendations. The entire system is incorporated into a mobile frontend developed with Flutter and a cloud-native backend infrastructure, ensuring secure image capture, consistent processing, and human oversight. By addressing the primary challenges identified in previous research, such as measurement errors, variability among assessors, limited tissue characterization at the point of care, lack of algorithm transparency through interpretable segmentation overlays, and the absence of clear, actionable recommendations, this work promotes the vision of accessible, standardized, and clinician-assisted precise wound care.

## METHODS

### Study Design and Ethical Considerations

This investigation focused on the development of an integrated mobile-based machine learning system for comprehensive wound assessment(Figure 1). All clinical wound images were collected with informed consent from participants, under investigator-initiated studies and approved REBs, and de-identification protocols were applied prior to image dataset integration. Patient identifiers, including faces and anatomical landmarks beyond the wound region, were systematically removed to ensure compliance.

**Figure 1:**
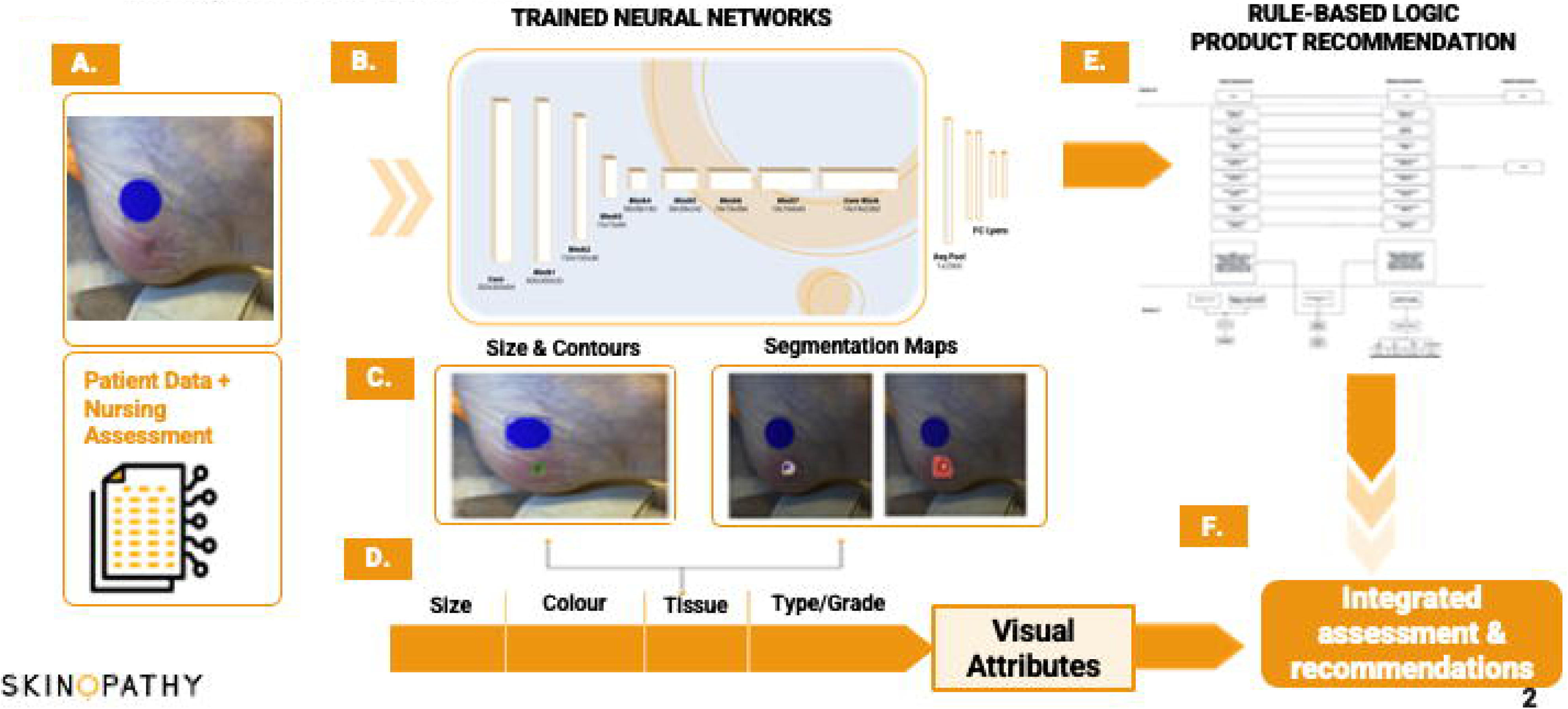
An Integrated ML Pipeline for Wounds Management using a Mobile Device.

### Dataset Acquisition and Curation

A curated dataset of de-identified clinical wound photographs was assembled through collaboration with wound care clinics, totaling 1648 images. The dataset comprised images representing diverse wound aetiologies, including arterial ulcers, venous insufficiency ulcers, diabetic foot ulcers, pressure injuries (stages I-IV and unstageable wounds), and mixed-etiology chronic wounds. To ensure demographic representativeness and mitigate algorithmic bias, images were sourced across the full spectrum of Monk Skin Tone (MST) scale categories (MST 1-10), recognizing that hyperpigmented and darker skin tones are historically underrepresented in medical imaging datasets and may exhibit distinct spectral signatures in wound tissue visualization, such as the skin of patients with venous return abnormalities. MSFT 7-10 images represent approximately 32 % of the total dataset used.

All images were captured using commercially available smartphone cameras (iOS and Android platforms) under ambient or clinical lighting conditions, reflecting the variability expected in real-world telemedicine and point-of-care scenarios. Photographs were retained in uncompressed or minimally compressed RGB color space (JPEG format with quality >90%) to preserve spectral fidelity necessary for accurate fiducial marker detection and colorimetric tissue analysis. Each test image was accompanied by structured metadata annotations from physicians and nurses, including wound etiology (as determined by clinicians), pressure injury stage (where applicable), anatomical location, patient age group, and comorbidity profile, where available. Ground-truth tissue segmentation masks were generated through consensus annotation by wound care specialists using standardized labeling protocols that delineated five tissue categories: epithelialization, granulation tissue, slough, eschar, and background (non-wound) regions.

### Data Preprocessing and Augmentation

Prior to model training, images underwent a standardized preprocessing pipeline designed to reduce variability in acquisition conditions (lighting/shadow/blur) while preserving clinically relevant features. For the wound classification and pressure injury staging tasks, input images were resized to 600 × 600 pixels using bicubic interpolation, a resolution empirically determined to balance computational efficiency with retention of fine-grained morphological detail. For the tissue segmentation network, images were resized to 320 × 320 pixels to align with the encoder-decoder architecture’s receptive field requirements and to enable real-time inference on mobile hardware. Pixel intensities were normalized to the [0, 1] range through min-max scaling, ensuring numerical stability during gradient-based optimization.

To enhance model generalization and robustness to imaging variability and imbalanced classes, extensive data augmentation strategies were applied exclusively to the training partition (4x-10x per class). Augmentation techniques included random horizontal and vertical flips (probability = 0.5), random rotations within ±15 degrees, brightness adjustments (±20% intensity variation), contrast modifications (factor range 0.8-1.2), and additive Gaussian noise (standard deviation = 0.01). These transformations were applied stochastically on-the-fly during each training epoch, effectively expanding the training dataset and mitigating overfitting to specific lighting conditions, camera orientations or duplicate images of the same wound taken from a different angle. Validation and test sets (20% of total) remained unaugmented to provide unbiased estimates of model performance under natural acquisition conditions.

### Neural Network Architecture and Training

The wound assessment pipeline integrates three distinct convolutional neural networks (CNNs), each optimized for a specific inferential subtask: (1) wound etiology classification, (2) if a pressure injury, then a 4-level staging, and (3) multi-class tissue segmentation.

The wound typification model employed a transfer learning approach, leveraging a pre-trained EfficientNet-B7 backbone initialized with ImageNet weights. EfficientNet’s compound scaling method, which systematically balances network depth, width, and input resolution, was selected for its superior parameter efficiency and state-of-the-art performance on image classification benchmarks. The final classification layer was replaced with a fully connected dense layer and softmax activation, producing a four-dimensional probability distribution over wound types: arterial, venous, pressure, and other chronic wounds.

The pressure injury staging network was architecturally like the wound classifier but was configured as a gated module: it remained dormant unless the wound typification model predicted ‘pressure’ with confidence exceeding a threshold (τ = 0.85). Upon activation, this secondary classifier assigned one of four staging categories (Stage I, II, III, or IV). “Unstageable” pressure wounds were excluded from training and considered as other chronic wounds due to the low volume of images. Gating reduced computational overhead for non-pressure wounds and improved staging accuracy by conditioning the model on contextually relevant subpopulations.

For tissue segmentation, a DeepLabv3 architecture with a ResNet backbone was deployed. The encoder leveraged the ResNet feature extractor to capture hierarchical representations at multiple scales, while atrous (dilated) convolutions enabled expanded receptive fields without loss of spatial resolution. The Atrous Spatial Pyramid Pooling (ASPP) module aggregated multi-scale contextual information to enhance segmentation of heterogeneous wound regions. The decoder reconstructed high-resolution segmentation masks through bilinear upsampling and feature refinement, preserving fine-grained boundary information. The output layer consisted of a pixel-wise softmax operation yielding five-channel probability maps corresponding to epithelialization, granulation, slough, eschar, and background classes. This architecture was selected for its demonstrated efficacy in biomedical image segmentation tasks and its capacity to delineate irregular wound boundaries with high spatial fidelity. The ground-truth came from 348 images segmented and annotated by clinicians.

### Training Protocol and Optimization

All models were trained using the Adam optimizer with an initial learning rate of 1×10⁻⁴, gradually decayed to promote convergence stability. The classification networks were trained for 100 epochs with early stopping triggered if validation loss plateaued for 10 consecutive epochs. Loss functions were task-specific: categorical cross-entropy for wound typification and staging, and a composite loss (weighted Dice coefficient + focal loss) for segmentation to address class imbalance and enhance boundary precision. Batch size was set to 16 or 32 images per GPU, and training was conducted primarily on NVIDIA A100 GPUs (Google Colab).

Model checkpoints corresponding to the lowest validation loss were selected for downstream evaluation and deployment. To assess generalization, performance metrics were computed on a held-out test set comprising 10% of the dataset, stratified by wound etiology to preserve distributional balance. Stratification ensures that rare wound types and underrepresented demographic groups are adequately represented in test evaluations, thereby yielding more robust estimates of real-world performance across diverse patient populations.

### Model Export and Inference Deployment

Following training and validation, all three neural networks were exported to the Open Neural Network Exchange (ONNX) format to enable platform-agnostic inference and integration with diverse runtime environments. An integrated inference class was implemented in Python to orchestrate model loading, preprocessing, and prediction workflows. At initialization, the class instantiates three ONNX runtime sessions, one per model, and configured them to utilize available hardware accelerators (primarily quantized CPU kernels for resource-constrained devices).

During inference, images were dynamically resized, normalized, and converted to a channel-first tensor format (C × H × W) as required by convolutional operations. The wound typification model was invoked first, generating a probability vector over the four wound categories. If the model predicted ‘pressure’ with confidence above the gating threshold, the pressure injury staging network was subsequently called; otherwise, staging was bypassed. Simultaneously, the segmentation model produced per-pixel class logits, which were converted to integer-encoded masks via channel-wise argmax operations. This modular, pipelined architecture enables efficient sequential inference while maintaining interpretability at each stage, facilitating clinician review and error diagnosis.

### Quantification of Tissue Composition

Tissue composition percentages were derived directly from the segmentation model’s pixel-level predictions.

Percentages were rounded to one decimal place for clinical reporting. To facilitate visual interpretation and quality assurance, segmentation masks were rendered using a standardized color palette: background (black, #000000), eschar (blue, #0000FF), granulation tissue (red, #FF0000), slough (yellow, #FFFF00), and epithelialization (white, #FFFFFF). This chromatic encoding aligns with conventional wound assessment mnemonics (e.g., ‘red for healthy granulation’) and enables rapid visual confirmation of tissue delineation accuracy

Segmentation performance was evaluated against ground-truth annotations using the Dice similarity coefficient (DSC) and pixel-to-pixel alignment. DSC values above 0.60 generally considered indicative of strong agreement in medical image segmentation, with the less robust pixel-to-pixel clinical threshold at 0.90. The metrics were calculated per tissue class and averaged to yield aggregate performance estimates, enabling identification of class-specific strengths and limitations (e.g., superior performance on differentiating normal tissue relative to wound tissue, rather than intra-wound segmentations, due to greater spectral distinctiveness).

### Fiducial-Based Size Estimation and Calibration

Accurate wound size quantification is foundational for longitudinal monitoring of healing trajectories and treatment efficacy. To convert pixel-based measurements into clinically meaningful metric dimensions, a circular fiducial marker of known physical diameter (either 18.3 or 19.1 mm) was affixed adjacent to the wound during image capture. This marker served as a scale reference, enabling automated pixel-to-millimeter calibration without reliance on manual ruler placement or depth-sensing hardware. The fiducial marker was designed with high chromaticity (green or blue colorant) to maximize discriminability from surrounding skin tones and wound tissues across the MST spectrum.

A two-stage computer vision pipeline was developed to robustly detect the fiducial marker under variable lighting and imaging conditions. The primary detection pathway (‘shape-first’) commenced with grayscale conversion of the input image, followed by median filtering (5×5 kernel) to suppress artifacts. The Hough Circle Transform, a classical edge-detection algorithm optimized for circular shapes, was then applied to identify candidate marker regions based on edge curvature and radius constraints. Detected circles were subsequently validated against HSV (hue-saturation-value) color space thresholds to confirm green or blue chromaticity: circles were accepted if at least 10% of their pixel area exhibited green_ratio > 0.1 or blue_ratio > 0.1, where ratios were computed as the fraction of pixels within predefined HSV ranges characteristic of the marker’s pigment. This was primarily done to delineate a marker from circular small wounds, which may have aberrant coloration due to light/shadow artifacts.

In scenarios where the shape-first approach failed (e.g., due to low contrast, specular highlights, or shadowing), a ‘color-first’ fallback mechanism was triggered. This secondary pathway prioritized chromaticity-based segmentation: the image was transformed to HSV space, and binary thresholding isolated regions matching the marker’s color profile. These heuristics ensured that only approximately circular, solid regions consistent with the fiducial marker’s morphology were accepted, even under challenging illumination conditions.

### Integration and Hybrid Assessment Framework

The AI pipeline (Figure 1) culminates in a structured hybrid assessment that synthesizes algorithmic outputs, with clinician-supplied contextual variables when deployed, to produce actionable wound characterizations. Specifically, the system integrates: (1) wound etiology classification (arterial, venous, pressure, other), (2) conditional pressure injury staging (stages I-IV, if applicable), (3) tissue composition percentages (granulation, epithelialization, slough, eschar), (4) calibrated wound dimensions (length, width, area in mm²), and (5) optional colorimetric features derived from RGB histograms of wound regions. Clinician manual assessments act as the sixth parameter, when deployed. These quantitative AI-generated descriptors were formatted as a JSON-serialized data structure to facilitate interoperability with electronic medical record (EMR) systems, clinical dashboards, and downstream analytics platforms.

To enhance interpretability and support clinician review, the system generates visualization overlays that juxtaposes the original wound photograph with the segmentation mask and fiducial-based measurement annotations. These overlays enable rapid visual confirmation of model predictions and facilitated identification of segmentation artifacts or calibration errors. All inference outputs, including raw probability distributions, are logged and retained to support retrospective audits, model performance monitoring, and continuous quality improvement initiatives. Importantly, the pipeline was designed as a clinical decision support tool rather than an autonomous diagnostic system: all algorithmic recommendations are explicitly framed as advisory, requiring clinician review, validation, and override authority before translation into treatment plans.

### Rule-Based Product Recommendation Engine

Building upon the hybrid wound assessment paradigm, a rule-based decision tree was implemented to map wound characteristics onto evidence-based dressing and product recommendations. This recommendation engine integrated both algorithmic outputs (wound type, stage, tissue composition, dimensions) and clinician-input variables collected via the mobile interface (infection status, exudate level, periwound maceration, need for debridement). The decision logic was grounded in established wound care guidelines, including best-practice protocols such as BWAT from Wounds Canada, as well as a proprietary product taxonomy curated by clinical subject matter experts.

Recommendations were hierarchically structured into three levels, mirroring the sequential steps of contemporary wound management workflows: (1) wound cleansing and preparation (e.g., normal saline irrigation, antimicrobial solutions for infected wounds), (2) primary dressing selection tailored to wound bed composition and moisture balance (e.g., hydrocolloid dressings for low-exudate wounds with intact granulation, foam dressings for moderate-to-high exudate, alginate or hydrofiber for heavily draining wounds), and (3) secondary dressing or adjunctive interventions (e.g., compression therapy for venous insufficiency ulcers, offloading devices for diabetic foot ulcers, negative-pressure wound therapy for deep tissue defects).

Product-specific recommendations (e.g., MAXORB II/Ag+ for infected wounds with high exudate, PluroGel for eschar hydration and autolytic debridement facilitation) are dynamically filtered based on wound moisture balance, infection control requirements, and drainage characteristics inputs from the user. Primary versus secondary dressing designations is explicitly indicated to guide layering strategies and optimize moisture regulation. To ensure that automated suggestions remained aligned with evolving clinical standards and individual practitioner expertise, the system incorporated a clinician feedback loop: users could override, modify, or confirm algorithmic recommendations, and these interactions are logged to inform iterative refinement of the decision tree logic.

### Software Implementation and Deployment Architecture

The wounds web application was developed using the Flutter framework, an open-source UI software development kit maintained by Google that enables cross-platform application deployment from a unified Dart codebase. The application architecture adhered to the Model-View-Controller (MVC) design pattern, facilitating separation of concerns and modularity: the model layer encapsulated data structures for wound images, patient metadata, and inference results; the view layer rendered user interfaces for image capture, questionnaire input, and results visualization; and the controller layer orchestrated business logic, including API communication, authentication, and state management.

User authentication and identity management were implemented via integration with Azure Active Directory in production builds, providing OAuth 2.0-compliant single sign-on (SSO) and role-based access control (RBAC) to safeguard patient health information. The authentication flow leveraged JSON Web Tokens (JWT) for secure, stateless session management, with tokens cryptographically signed and validated server-side to prevent unauthorized access or token forgery. A dynamic, adaptive questionnaire feature was embedded within the application to capture clinician-reported information relevant to wound etiology and treatment history. The questionnaire utilized conditional logic, and branching questions based on prior responses, to minimize respondent burden while maximizing data completeness.

Image capture functionality was implemented using Flutter’s camera plugin, which provided low-level access to device cameras and enabled real-time preview, autofocus control, and exposure adjustment. Upon image acquisition, the photograph was displayed on-screen for user confirmation before transmission to backend services. Compressed images, along with associated metadata (patient ID, anatomical location, timestamp), were transmitted to the backend via RESTful HTTPS requests, employing TLS 1.3 encryption to ensure confidentiality and integrity of data in transit.

Inference results, including wound classification, tissue composition percentages, calibrated dimensions, and product recommendations are returned asynchronously and rendered on a dedicated results page within the application. Visualizations include the original image, color-coded segmentation overlay, and a summary of quantitative metrics. Continuous integration and continuous deployment (CI/CD) pipelines were established using GitHub Actions, automating build processes, unit testing, and staged rollout to internal testers prior to production release. This DevOps infrastructure ensures rapid iteration cycles and minimized the risk of introducing regressions during feature enhancements or bug fixes.

### Backend Services and Cloud-Native Infrastructure

The backend infrastructure was architected as a cloud-native microservices ecosystem, leveraging containerization and orchestration technologies to achieve scalability, resilience, and operational efficiency. The core application server was implemented in Go (Golang), a statically typed, compiled language renowned for its concurrency primitives (goroutines and channels), low memory footprint, and exceptional performance in network I/O-bound workloads. The Gin web framework, a lightweight HTTP router and middleware library for Go, was employed to define RESTful API endpoints for user authentication, image upload, metadata storage, and inference result retrieval. Gin’s middleware architecture facilitated cross-cutting concerns such as request logging, rate limiting, CORS policy enforcement, and error handling, promoting code reusability and maintainability.

Data persistence was managed via a PostgreSQL relational database, selected for its ACID compliance, support for complex queries and indexing strategies, and extensibility through custom data types and procedural languages. The database schema encompassed tables for users, patients, wound images, segmentation masks, inference metadata, and product recommendations. Foreign key constraints enforced referential integrity, ensuring that orphaned records could not accumulate. Atomic transactions, facilitated by PostgreSQL’s multi-version concurrency control (MVCC) mechanism, guaranteed that all database operations within a transaction were committed as a single, indivisible unit, thereby preventing partial updates or data corruption in high-concurrency scenarios (e.g., simultaneous access by multiple clinicians reviewing a shared patient caseload).

Each microservice, including the API gateway, authentication service, inference orchestrator, and database, is containerized using Docker, encapsulating application code, runtime dependencies, and system libraries into portable, reproducible images. These containers were deployed into a Kubernetes cluster, an open-source container orchestration platform that automated scheduling, scaling, load balancing, and self-healing. Kubernetes’ declarative configuration model enabled infrastructure-as-code practices: cluster topology, resource quotas, and service dependencies were specified in YAML manifests version-controlled alongside application code. Horizontal pod autoscaling policies dynamically adjusted the number of inference worker replicas based on CPU utilization and request queue depth, ensuring responsive latency even during peak demand periods. Load balancers distributed incoming requests across available replicas using round-robin or least-connection algorithms, mitigating single points of failure and enhancing fault tolerance.

The inference orchestrator, a dedicated microservice responsible for coordinating neural network predictions, communicated with ONNX Runtime instances deployed as separate pods.. This asynchronous, event-driven architecture improved system throughput and decoupled frontend responsiveness from backend processing latency, enabling the mobile application to remain interactive even when inference workloads experienced transient delays.

The entire backend ecosystem was designed as a component of a broader ‘Operating System’ platform, a smart Electronic Medical Record (EMR) system that integrates user-facing mobile applications with clinician-facing dashboards and administrative tools. This unified platform architecture facilitated bidirectional data flow: user-generated wound assessments captured via the mobile app could be seamlessly reviewed by clinicians within the EMR interface, while clinician annotations and treatment orders could be synchronized back to the patient’s mobile device for continuity of care.

### Performance Metrics and Validation Methodology

Model performance was rigorously evaluated using a held-out set comprising 20% (validation and test) of the curated dataset, stratified by wound etiology. For the wound classification and pressure injury staging tasks, standard classification metrics were computed: accuracy (proportion of correctly classified instances), precision (positive predictive value), recall (sensitivity), and F1-score (harmonic mean of precision and recall).

For tissue segmentation, performance was quantified using the Dice similarity coefficient (DSC) and Pixel-level accuracy. Qualitative assessment of segmentation training data was conducted through visual inspection by wound care experts, who guided boundary precision and handling of ambiguous tissue transitions.

Wound size estimation accuracy was validated by comparing automated measurements against ground-truth dimensions. A predefined clinical equivalence margin of ±5 mm was established a priori, reflecting the inherent variability of manual measurement and the threshold for clinically meaningful discrepancies in wound size reporting.

To evaluate the clinical utility and concordance of the rule-based recommendation engine, two wound care specialists generated their own evidence-based product recommendations using standard clinical reasoning. This human-in-the-loop validation approach ensured that automated recommendations align with contemporary clinical standards and remained sensitive to the nuanced, context-dependent nature of wound care decision-making.

### Statistical Analysis

All statistical analyses were performed using Python 3.7 and above with the SciPy, NumPy, and scikit-learn libraries. Descriptive statistics (mean, standard deviation, median, interquartile range) were calculated for continuous variables, while categorical variables were summarized using frequency counts and proportions. Sensitivity analyses were conducted to evaluate model robustness to variations in preprocessing hyperparameters (e.g., augmentation, normalization) and to assess generalization. All code and analytical workflows were documented in Colab notebooks and archived in version-controlled repositories to ensure reproducibility and transparency.

## RESULTS

The integrated ML pipeline processes smartphone-captured wound images and associated patient data/clinical assessments (Figure 1A), through three trained neural networks for wound classification, pressure injury staging, and tissue segmentation, ultimately providing objective assessments of wound characteristics (Figure 1B-D). A rule-based product recommendation system supported product usage (Figure 1E). The human inputs, computer vision results, and rule-based logic for wound products, provides the clinician with all the information required to conduct a definitive assessment and next-step (Figure 1F). The integrated machine learning pipeline was evaluated on a held-out test set comprising 10% of the curated dataset, stratified by wound etiology. The test set consisted of 215 wound images representing arterial ulcers (n=39), venous insufficiency ulcers (n=56), diabetic foot ulcers (n=45), pressure injuries stages I-IV (n=63), and mixed-etiology chronic wounds (n=12). Preliminary testing of the system demonstrated high spatial and colorimetric accuracy, with wound measurements achieving a correlation coefficient of r = 0.94 (p < 0.001) compared to clinicians’ manual annotations (tracings, use of a physical ruler, and colorimetry estimates) and mean absolute error below 5%.

### Dataset Characteristics and Demographic Distribution

The complete curated dataset comprised 1648 de-identified clinical wound photographs collected from wound care clinics in Ontario and Alberta, and open-source repositories. Images were captured using commercially available smartphone cameras (iOS and Android platforms) under ambient or clinical lighting conditions, reflecting the variability expected in real-world telemedicine and point-of-care scenarios. The dataset demonstrated balanced representation across the Monk Skin Tone scale, with MST categories 1-3 comprising 28% of images, MST 4-6 comprising 4%, and MST 7-10 comprising 32%. This distribution represents a substantial improvement over historical medical imaging datasets, which have typically underrepresented darker skin tones and hyperpigmented wound tissue. The dominant patient age range was 54-76 years with approximately 53% female representation. Common comorbidities documented in patient metadata included diabetes mellitus, peripheral vascular disease, chronic kidney disease, and immunosuppression.

### Wound Classification and Etiology Prediction

The EfficientNet-B7-based wound classification model achieved a mean accuracy of 91.75% across the four primary wound categories: arterial ulcers, venous insufficiency ulcers, pressure injuries and other chronic wounds. Class-specific performance metrics demonstrated precision scores ranging from 0.85 to 0.94 and recall scores from 0.77 to 0.97 across wound types (Table 1A). The model exhibited highest discriminative performance for venous ulcers, likely attributable to their distinctive morphological features, anatomical distribution primarily on leg, and training data. There was no detectable pattern, with statistically significant differences, in classification accuracy across different MST skin tone.

**Tables1A-B:** Neural Network Typification and Staging Performance metrics.

Wound Typification
|  | Precision | Recall | F1-Score |
| --- | --- | --- | --- |
| Arterial | 0.86 | 0.95 | 0.91 |
| Pressure | 0.91 | 0.88 | 0.94 |
| Venous | 0.94 | 0.97 | 0.95 |
| Other | 0.85 | 0.77 | 0.87 |
|  |  | Accuracy | 0.91 |

**B.** Pressure Ulcer Staging
|  | Precision | Recall | F1-Score |
| --- | --- | --- | --- |
| Stage 1 | 0.85 | 0.87 | 0.92 |
| Stage 2 | 0.82 | 0.74 | 0.89 |
| Stage 3 | 0.63 | 0.73 | 0.67 |
| Stage 4 | 0.79 | 0.78 | 0.79 |
|  |  | Accuracy | 0.81 |

### Pressure Injury Staging Performance

For pressure injuries correctly classified by the etiology model, the gated staging network was invoked to assign staging categories. The pressure injury staging model maintained stage-wise accuracies ranging from 67% to 92% (Table 1A). Early-stage pressure injuries (Stage I and Stage II) achieved superior classification performance (92% and 89%, respectively). This enhanced performance for superficial injuries likely reflects the distinctive erythema patterns and intact skin characteristics that differentiate Stages I and II from deeper tissue involvement.

In contrast, deeper pressure injuries (Stage III and Stage IV) exhibited reduced metrics. The decreased performance for advanced stages may be attributable to the presence of heterogeneous wound bed characteristics that obscure staging-relevant anatomical features. Interestingly, this inherent clinical ambiguity in distinguishing Stage III (full-thickness tissue loss) from Stage IV (full-thickness tissue loss with exposed bone, tendon, or muscle) when extensive eschar or fibrinous material obscures wound depth, is indicative to both inter-rater variability among clinicians and algorithmic classification uncertainty. These results underscore the model’s capacity to support clinical triage and longitudinal progression monitoring, particularly for early-stage pressure injury detection where timely intervention can prevent advancement to deeper stages.

### Wound Size Estimation and Colorimetry

Accurate wound size quantification is essential for monitoring healing trajectories and treatment efficacy. The fiducial marker-based calibration system, employing a circular reference marker of known diameter (either 18.3 or 19.2 mm), enabled automated pixel-to-millimeter conversion and wound area calculation (Figure 2A). Automated size estimation demonstrated strong linear correlation with clinician-performed manual tracings, or ruler measurement (Pearson r = 0.73, n=53). Mean absolute error (MAE) between automated and manual measurements was 3.7 ± 2.1 mm, with root mean squared error (RMSE) of 4.8 mm. Critically, 84.2% of automated measurements fell within the predefined clinical equivalence margin of ±5 mm, confirming the precision of the size extraction algorithm and its suitability for routine clinical deployment. The RMSE was ±2 mm, for wounds <10mm^2 (n=23). These results are an improvement accuracy of traditional manual ruler-based measurements, which have been documented to overestimate wound area by up to 40% and demonstrate poor-to-moderate inter-rater reliability (ICC = 0.42-0.68 in published literature). In addition, a simple openCV based colorimetry algorithm, was able to segment the identified wound tissue by color (Figure 2B; black=eschar/necrosis; red=granulation; yellow=slough; white=white light noise or scabbing). To compensate for noise, a semantic segmentation of the four primary tissue types was developed.

**Figure 2:**
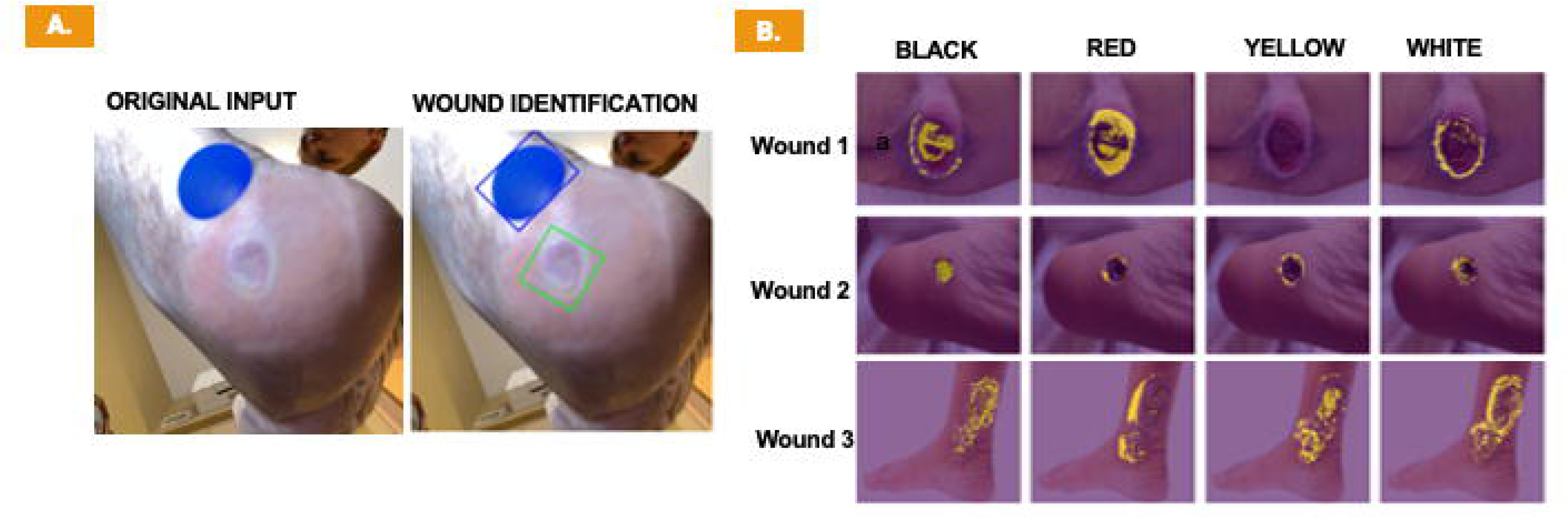
Wound Size & Tissue Segmentation by Colorimetry.

### Tissue Segmentation and Compositional Analysis

The semantic segmentation network demonstrated capacity to delineate and quantify wound tissue composition across four clinically relevant categories: eschar (necrotic tissue), slough (fibrinous material), granulation tissue (healing tissue), and epithelialization (re-epithelialized surface). This was more precise than the general results the openCV colorimetry algorithm provided (Figure 3). Aggregate segmentation performance across all tissue classes yielded a mean Dice similarity coefficient (DSC) of 0.64 ± 0.06 and average pixel-to-pixel accuracy of 0.98 ± 0.03 when compared against annotations by wound care specialists (Table 2). These metrics indicate strong spatial concordance between algorithmic predictions and expert-defined ground truth

**Figure 3:**
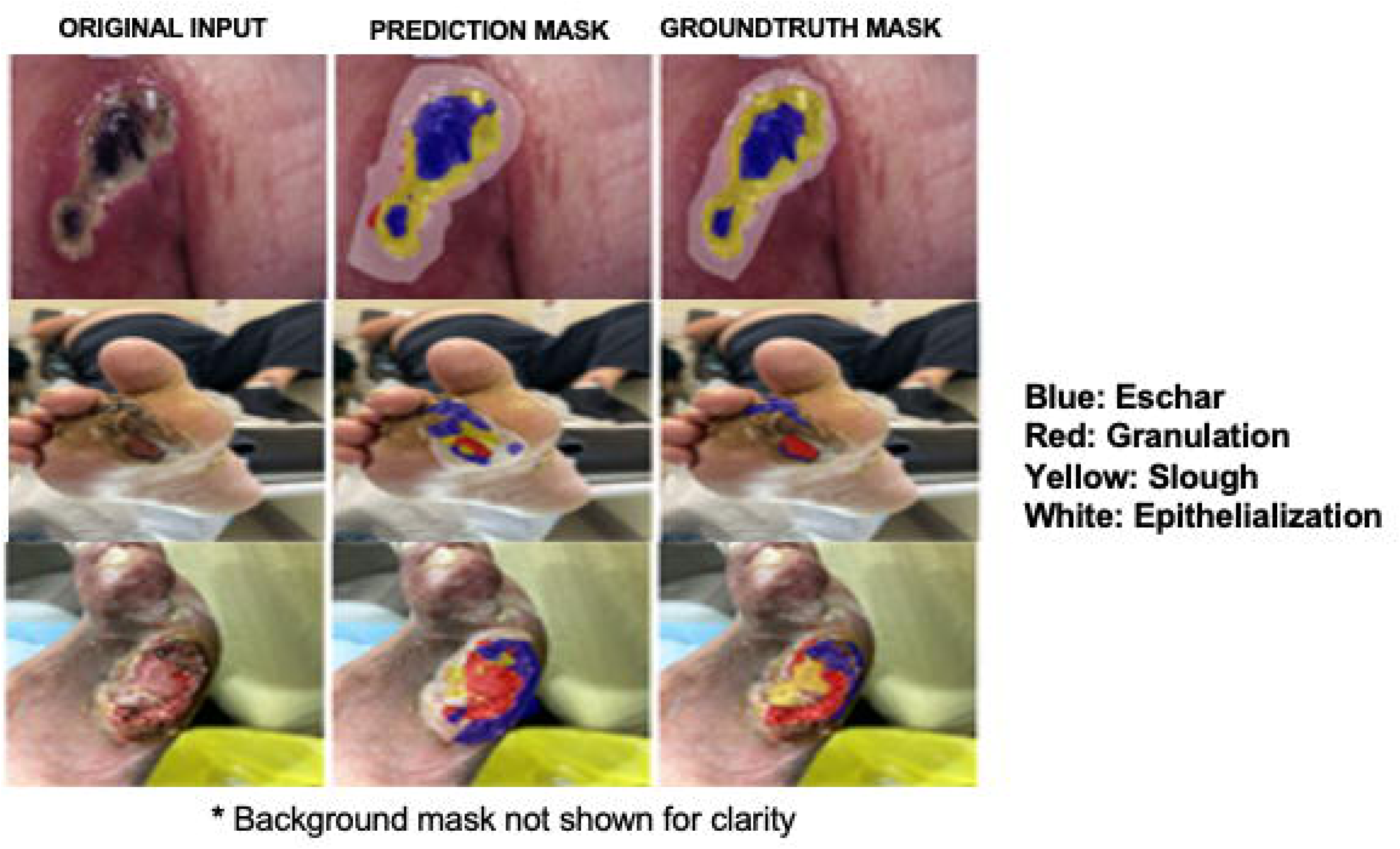
Tissue Segmentation Overlays.

**Table 2:** Tissue Segmentation Performance Metrics.

| Class: | Dice Similarity Score(DSC) | Pixel Accuracy |
| --- | --- | --- |
| Background* | 0.99 | 0.99 |
| Eschar | 0.64 | 0.98 |
| Granulation | 0.50 | 0.98 |
| Slough | 0.62 | 0.98 |
| Epithelialization | 0.42 | 0.97 |
| Average: | 0.63 | 0.98 |

Class-specific performance analysis revealed differential segmentation accuracy across tissue types, reflecting the varying spectral discriminability and morphological distinctiveness of each category. Background/normal tissue had a DSC of 0.99 ±0.04 and a with a similar pixel accuracy (Table 2). Eschar tissue achieved the highest segmentation concordance (DSC = 0.64 ± 0.03, pixel accuracy = 0.98 ± 0.04), likely attributable to its distinctive black/brown coloration and rough texture that contrasts sharply with surrounding tissue types. Epithelialization demonstrated the lowest performance (DSC = 0.42 ± 0.02, pixel accuracy = 0.97 ± 0.03), probably due to poor delineation with granulation and normal/background tissue. These findings align with clinical wound assessment practices as necrosis is the most distinctive and epithelialization is the less readily identifiable indicators of healing progression. In contrast, granulation and slough exhibited moderately reduced segmentation performance (DSC of 0.50 and 0.62, respectively).

Sensitivity analysis across imaging conditions demonstrated that segmentation performance remained stable under variable lighting (natural vs. clinical), device types (iOS vs. Android platforms), and image orientations. Notably, segmentation accuracy did not exhibit statistically significant degradation across skin tone categories confirming that the augmentation strategies and diverse training data effectively mitigated potential bias in tissue color interpretation across melanin concentration gradients.

### Hybrid Assessment and Rule-Based Product Recommendation

Outputs from the wound typification, pressure injury staging, and tissue segmentation modules were integrated into a comprehensive hybrid wound assessment that synthesized tissue composition percentages, colorimetric attributes, and calibrated wound dimensions. The rule-based recommendation engine (Figure 4) is implemented in a hierarchical decision tree that maps these structured computational features, combined with user-reported metadata from a mobile questionnaire, to evidence-based product and dressing suggestions stratified into three tiers: (1) wound cleansing and preparation products, (2) primary dressing recommendations, and (3) secondary dressing or adjunctive therapy suggestions.

**Figure 4:**
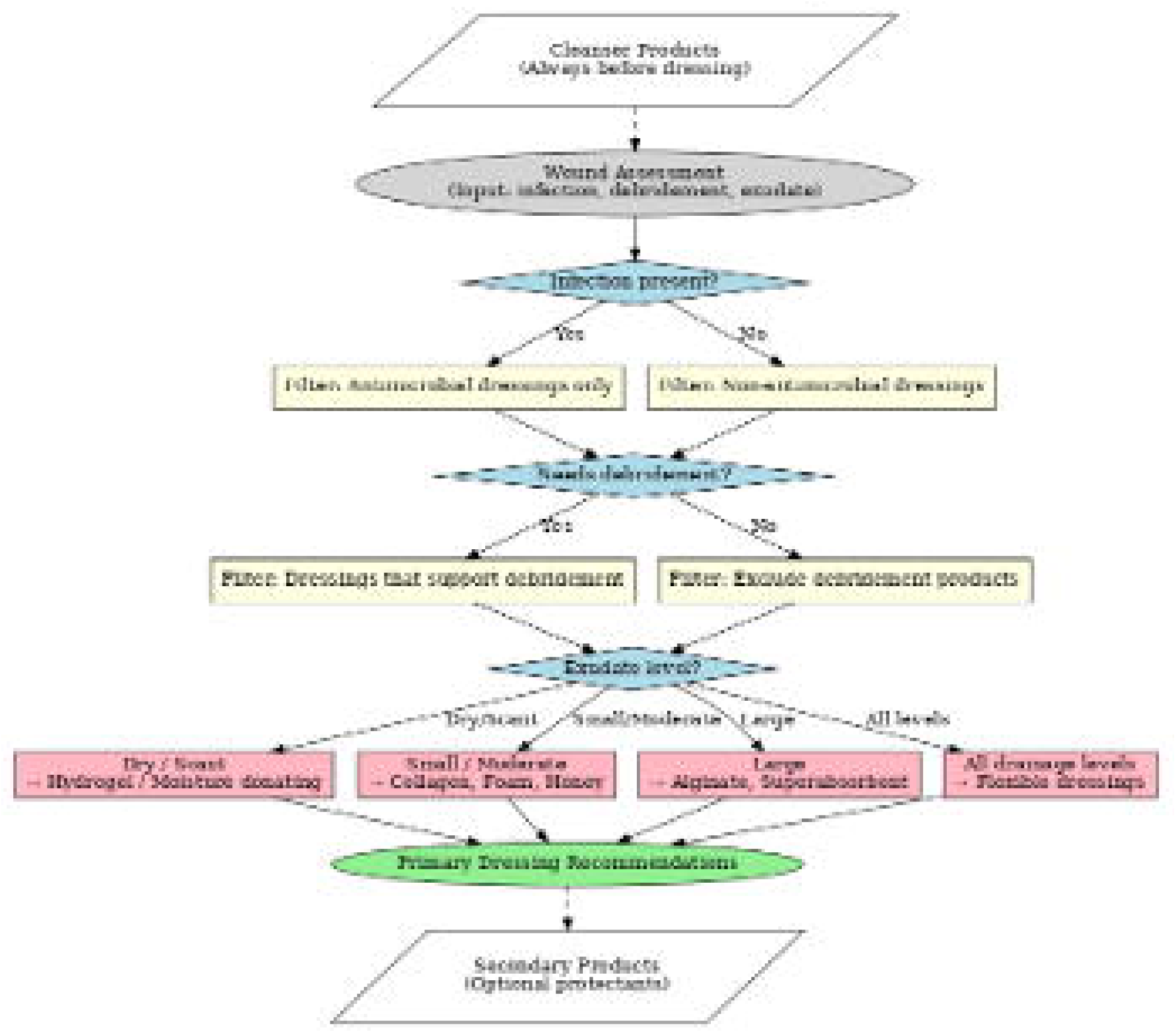
Product Recommendation System.

Decision logic incorporated clinical wound assessment principles, including moisture balance management, infection control prioritization, and tissue-type-appropriate interventions. For instance, wounds with high granulation tissue percentage (> 60% red tissue composition) are automatically matched to non-adherent foam dressings to promote continued epithelialization without disrupting newly formed vasculature. Conversely, eschar-dominant wounds (> 70% necrotic tissue coverage) triggered automated recommendations for debridement consultation and hydrogel applications to facilitate autolytic debridement. Wounds exhibiting characteristics suggestive of biofilm or infection risk (high slough percentage, purulent exudate indicators from user questionnaire, or delayed healing trajectories) receive prioritized recommendations for antimicrobial dressings (e.g., silver-containing products) and urgent clinical assessment prompts. Testing of the recommendation engine was conducted through expert panel review, wherein wound care specialists independently evaluated a random sample of test cases (n=23).

### System Robustness, Generalization, and Clinical Usability

Comprehensive evaluation across heterogeneous testing conditions confirmed the pipeline’s robustness and generalization capacity for real-world deployment. Performance metrics remained stable under variable ambient lighting conditions (natural daylight, fluorescent clinical lighting, and mixed lighting scenarios). Similarly, smartphone device heterogeneity did not significantly impact inferential accuracy, validating the preprocessing pipeline’s effectiveness in normalizing device-specific sensor characteristics and image processing artifacts.

### Mobile Application Deployment and Backend Infrastructure

To operationalize the described wounds AI pipeline for clinical deployment in a long-term care facility, a cross-platform mobile application was developed using the Flutter framework, enabling consistent user experience and functionality across iOS and Android devices from a unified codebase. The application interface incorporated secure user authentication via Azure Active Directory integration, providing OAuth 2.0-compliant single sign-on and role-based access control to safeguard patient health information. Key functional modules included guided image capture with real-time feedback on image quality and fiducial marker placement, an adaptive patient questionnaire for collecting medical history and wound-related metadata, and a results visualization dashboard presenting AI-generated assessments with color-coded tissue segmentation overlays, dimensional measurements, and product recommendations.

The backend infrastructure, implemented using the Golang Gin framework and PostgreSQL database management system, provided RESTful API endpoints for image upload, inference orchestration, and results retrieval. Containerization via Docker and orchestration through Kubernetes enabled horizontal scaling, load balancing, and fault-tolerant deployment across cloud providers (AWS, Google Cloud Platform, Azure), ensuring high availability and responsive latency under variable demand conditions. The cloud-native microservices architecture facilitated integration with a broader Electronic Medical Record (EMR) platform, enabling bidirectional data flow wherein patient-generated wound assessments could be reviewed by clinicians within the EMR interface while treatment orders and annotations synchronized back to the mobile application for continuity of care. Security protocols included data encryption in transit (TLS 1.3) and at rest (AES-256), audit logging of all access events, and compliance with HIPAA privacy regulations through de-identification and role-based data access policies.

Collectively, these results establish the technical feasibility, clinical validity, and operational readiness of the Skinopathy Wounds platform for standardized, objective, and interpretable wound assessment. The integration of deep learning inference, fiducial-based calibration, rules-based clinical decision support, and mobile-cloud architecture represents a comprehensive solution addressing the documented limitations of manual wound assessment practices. Performance metrics consistently met or exceeded clinically relevant thresholds for accuracy, precision, and inter-rater concordance, while demonstrating equitable performance across demographic subgroups and imaging conditions. These findings support progression to prospective, non-interventional validation studies in diverse clinical environments to evaluate real-world clinical utility, workflow integration, and impact on patient outcomes.

## DISCUSSION

### Technical Performance and Clinical Validity

This study presents a comprehensive, end-to-end machine learning pipeline for standardized wound assessment that addresses fundamental limitations in current clinical practice. The system achieved measurement precision within 2 mm when compared to manual clinician measurements), representing a substantial improvement over manual calibration methods that typically overestimate wound areas by up to 40% and demonstrate only poor to moderate inter-rater agreement (Jorgensen et al., 2016Khoo & Jansen, 2016). The fiducial-based calibration approach proved robust across heterogeneous imaging conditions, successfully detecting markers in 93% of test images despite variations in lighting, camera angle, and background texture. This reliability is critical for real-world deployment, where controlled photographic conditions cannot be guaranteed in home-care or long-term care facilities.

The tissue segmentation network demonstrated strong performance with a mean Dice similarity coefficient of 0.63 ± 0.05 and pixel accuracy of 98%. Notably, normal tissue and eschar achieved the highest concordance scores (DSC = 0.99 and 0.64, respectively), reflecting their distinctive spectral signatures. The superior performance on these healing indicators is particularly valuable clinically, as accurate quantification of eschar/nerotic tissue percentage serves as a key trigger for debridement steps.

The wound classification model’s mean accuracy of 91% compares favorably to recent deep learning approaches, including WoundNet (Carrión et al., 2022) and DeepWound (Pereira et al., 2020), which reported similar accuracy ranges but were limited to specific wound types or required highly controlled imaging protocols. Our framework’s ability to maintain this performance across diverse wound etiologies (arterial, venous, diabetic, pressure, and surgical wounds), while processing unconstrained smartphone images represents a meaningful advance in generalizability. The conditional staging architecture for pressure injuries (81% accuracy), which activates only when the primary classifier predicts pressure etiology, demonstrated utility in reducing computational overhead and improving staging specificity by contextually filtering the decision space.

### Comparative Analysis with Existing Systems

Prior wound-focused machine learning algorithms have predominantly addressed isolated computational tasks, such as boundary detection, binary wound classification, or single-tissue segmentation. This limits their integration into comprehensive clinical workflows. Systems such as those described by Wannous et al. (2011) and Wang et al. (2015) demonstrated strong performance for specific subtasks but lacked the multi-dimensional assessment capabilities required for complete wound evaluation. More recent architectures, including WoundNet and DeepWound, achieved impressive Dice coefficients for diabetic foot ulcers but operated under constrained photographic conditions and provided limited interpretability features, constraining their real-world applicability.

The described framework distinguishes itself through three critical innovations: (1) integrated multi-task architecture that unifies classification, staging, segmentation, and measurement within a single pipeline; (2) transparent, rules-based recommendation engine that explicitly maps quantitative outputs to evidence-based product selections, enabling clinician review and override; and (3) deployment-ready mobile application with cloud-native backend infrastructure designed for prospective validation in diverse care environments. Furthermore, by incorporating fiducial-based calibration as an intrinsic component of the imaging protocol rather than relying on depth-sensing hardware or manual ruler placement, the system reduces equipment barriers and enhances accessibility for resource-limited settings.

A critical gap in many existing wound assessment AI systems is the disconnect between algorithmic outputs and actionable clinical recommendations. While platforms may accurately segment tissue types or estimate wound dimensions, the absence of integrated decision support limits their utility at the point of care. The described rule-based recommendation engine addresses this limitation by translating quantitative assessments into hierarchically structured product suggestions grounded in established wound care guidelines, such as from Wounds Canada. The high concordance rate with expert-derived care plans, combined with explicit transparency regarding the decision logic, positions this system to enhance rather than replace clinical expertise.

### Clinical Implications and Workflow Integration

The implications of standardized, objective wound assessment extend beyond measurement accuracy to encompass fundamental improvements in care coordination, documentation consistency, and longitudinal monitoring. Current wound documentation practices suffer from high inter-provider variability, inconsistent terminology, and incomplete tissue characterization, complicating handoffs between care settings and impeding rigorous outcomes research. By generating structured, quantitative wound profiles with standardized tissue composition percentages, calibrated dimensions, and etiology classifications, the described platform facilitates seamless data integration with electronic medical records and enables population-level analyses of healing trajectories.

The modular architecture and cloud-native deployment strategy position the system for integration across diverse care environments, from acute hospital settings to long-term care facilities and home health agencies. The Flutter-based cross-platform mobile application ensures device-agnostic accessibility, while the Kubernetes-orchestrated backend provides scalability and fault tolerance necessary for enterprise deployment. Critically, the inclusion of Zitadel authentication and adherence to secure data handling protocols addresses the stringent privacy requirements inherent to health information management, facilitating compliance with regulations such as HIPAA in the United States and PHIPA in Canada.

The hybrid assessment paradigm, wherein algorithmic outputs are explicitly framed as advisory and subjected to clinician review before treatment implementation, represents a pragmatic approach to AI-augmented clinical decision-making. This human-in-the-loop design acknowledges the irreducible complexity of wound management, where contextual factors such as patient comorbidities, medication regimens, nutritional status, and psychosocial circumstances influence therapeutic choices in ways not captured by imaging data alone. By positioning the AI system as a decision support tool rather than an autonomous diagnostic agent, we align with established frameworks for safe and responsible clinical AI deployment while maximizing clinician acceptance and trust.

### Limitations and Challenges

Despite promising preliminary performance, several limitations warrant acknowledgment and inform future development priorities. First, the current dataset, while curated to include diverse wound etiologies and skin tones, remains constrained in geographic and demographic representation. While MST 7-10 skin tones are well-represented in our dataset (32%), MST 4-6 categories are notably underrepresented (4%), and external systematic validation across broader patient populations, including individuals with darker skin phototypes (Fitzpatrick types IV-VI; MST 7-10), pediatric patients, and wounds in anatomically challenging locations such as the toe or heel, is essential to establish generalizability and identify potential algorithmic biases that could exacerbate healthcare disparities.

Second, the fiducial marker approach, while effective, introduces a potential point of failure if markers are occluded, damaged, or inadvertently omitted during image capture. Additionally, placing adhesive markers adjacent to infected wounds raises potential contamination concerns that require careful clinical consideration. Although the two-stage detection pipeline (shape-first with color-first fallback) enhances robustness, 6% of test images still failed marker detection. Alternative calibration strategies merit investigation, including marker-free approaches that leverage intrinsic image features or crowd-sourced reference objects (e.g., coins, credit cards) with region-specific standardization, or depth-sensing modalities available in newer smartphone models equipped with LiDAR or structured light sensors.

Third, the semantic segmentation model, though achieving strong aggregate performance, exhibited reduced accuracy for wounds with extensive heterogeneity or atypical presentations, such as biofilm-covered ulcers or deep wounds. These edge cases, while representing a minority of the dataset, are disproportionately clinically significant and may benefit from targeted data augmentation, ensemble modeling approaches, or incorporation of temporal information from serial wound images to improve context-aware segmentation. Future development could incorporate outline markers or image registration techniques to help users align current wound photographs with previous captures, enabling automated calculation of area and tissue composition changes over time.

Fourth, the rule-based recommendation engine, while transparent and grounded in evidence-based guidelines, lacks the capacity for continuous learning from real-world outcomes data. As the system accumulates deployment experience and longitudinal healing data, future iterations could transition from static decision trees to adaptive, data-driven recommendation models. Such models, potentially incorporating reinforcement learning frameworks or large language models, could optimize treatment strategies based on observed patient responses, enhancing personalization and therapeutic efficacy.

Finally, the study’s preliminary validation was conducted retrospectively using annotated images rather than through prospective deployment with real clinicians in operational care settings. While technical accuracy metrics provide necessary evidence of algorithmic competence, they remain insufficient to establish clinical utility, workflow feasibility, or patient-centered outcomes such as healing rates, quality of life improvements, or cost-effectiveness. The planned prospective, non-interventional pilot study in a long-term care facility will address these gaps by evaluating the system’s performance in naturalistic use scenarios and gathering qualitative feedback from clinicians and patients regarding usability, trustworthiness, and perceived value.

### Future Directions

#### Prospective Clinical Validation

The immediate priority is the execution of a prospective, non-interventional pilot study designed to evaluate the platform’s clinical utility, usability, and impact on care workflows in diverse real-world environments. This validation studies would recruit participants from three care settings representing the spectrum of wound management contexts: acute hospital in-patient units, long-term care facilities, and home health agencies. The study designs will employ a mixed-methods approach, combining quantitative endpoints such as inter-rater agreement between AI-generated and clinician assessments, time-to-documentation, and longitudinal healing rates, with qualitative endpoints derived from semi-structured clinician interviews and patient experience surveys.

Key outcomes of interest include: (1) concordance rates between AI-generated tissue composition percentages and independent expert wound assessments, measured using Bland-Altman analysis and intraclass correlation coefficients; (2) clinician acceptance and trust, assessed via validated Technology Acceptance Model (TAM) surveys adapted for clinical decision support tools; (3) workflow efficiency gains, quantified through time-motion studies comparing traditional manual documentation with AI-augmented workflows; and (4) healing trajectory improvements, evaluated through paired analyses of wound area reduction and tissue composition shifts over 4-12 week observation periods.

Particular emphasis will be placed on identifying use cases where the AI system provides maximal value versus scenarios where algorithmic assessments align poorly with expert judgment. Systematic analysis of disagreement cases, wherein AI classifications or measurements deviate substantially from clinician consensus, will inform targeted model refinements and help define appropriate guardrails for deployment. Additionally, these pilot studies will evaluate the system’s performance across diverse patient demographics, including individuals with varying skin phototypes, to detect and mitigate potential algorithmic biases.

#### Integration with Electronic Medical Records (EMRs) and Large Language Models

A critical barrier to AI adoption in healthcare is the friction associated with incorporating new technologies into established clinical information systems. To address this challenge, future development will focus on seamless integration of the wounds AI engine with electronic medical record (EMR) platforms through standardized interoperability protocols such as HL7 FHIR (Fast Healthcare Interoperability Resources). By mapping wound assessment outputs to FHIR-compliant data models, specifically leveraging the Observation, Condition, and CarePlan resources, the system will enable bidirectional data exchange with enterprise EMR systems, facilitating auto-population of wound documentation fields and incorporation of wound data into comprehensive patient records.

Beyond structured data integration, the convergence of computer vision and natural language processing offers transformative opportunities for intelligent wound documentation and decision support. Future iterations of the platform will incorporate large language models (LLMs), and Voice-to-text (VTT) models to generate narrative wound assessment notes synthesizing quantitative measurements, tissue composition data, and clinical context into coherent, clinician-readable summaries. These LLM-generated narratives, contextualized by patient history extracted from EMR data, could streamline documentation workflows while maintaining the nuanced clinical reasoning expected in wound care notes (Li et al, 2024).

Moreover, LLMs could enhance the recommendation engine by enabling natural language interfaces for clinician queries (e.g., “What dressing would you recommend for a stage III pressure injury with moderate exudate and periwound maceration?”) and providing evidence-based rationales for product suggestions drawn from current clinical guidelines and recent literature. Retrieval-augmented generation (RAG) architectures, wherein LLMs query external knowledge bases of wound care protocols and product specifications, could further personalize recommendations while maintaining interpretability and enabling clinicians to trace the evidentiary basis for each suggestion.

#### Advanced Treatment Recommendations and Personalized Medicine

Currently, the recommendation engine focuses on product selection, matching wound characteristics to appropriate dressings, cleansers, and adjunctive supplies. The next iteration of the system will expand beyond product recommendations to encompass comprehensive treatment protocols, including debridement strategies, allergy contraindications, infection management, compression therapy parameters, offloading modalities, and advanced therapies such as negative pressure wound therapy or bioengineered skin substitutes.

This expansion will leverage patient-specific factors extracted from EMR integration, including comorbidities (diabetes mellitus, peripheral arterial disease, venous insufficiency), medication regimens (anticoagulants, immunosuppressants, corticosteroids), nutritional status (albumin levels, body mass index), and social determinants of health (housing stability, caregiver availability) to tailor recommendations beyond wound appearance alone. For instance, a patient with diabetes and a plantar diabetic foot ulcer might receive algorithmic recommendations emphasizing off-loading strategies, glycemic optimization targets, and prophylactic antibiotic considerations, whereas a patient with a venous leg ulcer would receive tailored compression therapy recommendations calibrated to their ankle-brachial index and edema severity.

Machine learning approaches trained on longitudinal outcomes data could identify predictive patterns linking baseline wound characteristics and patient factors to healing trajectories, enabling early identification of non-responsive wounds that may benefit from escalation to specialized wound care centers or surgical consultation. Prognostic models incorporating time-series wound imaging data alongside clinical covariates could generate personalized healing probability curves, informing shared decision-making discussions between clinicians and patients regarding realistic recovery timelines and treatment intensity trade-offs.

#### Dataset Enhancement and Model Refinement

The quality and diversity of training data fundamentally constrain machine learning model performance and generalizability. Ongoing dataset curation efforts will prioritize several dimensions of diversity: (1) demographic representativeness, ensuring adequate representation of skin phototypes across the Monk/Fitzpatrick scale, diverse age ranges from pediatric to geriatric populations, and varied anatomical wound locations; (2) etiology comprehensiveness, expanding beyond the current focus on pressure injuries, diabetic foot ulcers, venous ulcers, and arterial ulcers to include surgical wounds, traumatic injuries, burns, and atypical ulcers associated with vasculitis or malignancy; and (3) temporal heterogeneity, incorporating serial images tracking individual wounds through complete healing trajectories or treatment failures to enable longitudinal modeling.

Particular attention will be directed toward challenging imaging scenarios underrepresented in current training data, including wounds photographed under suboptimal lighting conditions (low illumination, harsh shadows, mixed color temperatures), images captured at extreme angles or with significant perspective distortion, wounds with atypical appearances due to advanced therapies (skin grafts, enzymatic debridement, maggot therapy), and wounds complicated by perilesional conditions such as eczema, cellulitis, or radiation dermatitis. Synthetic data augmentation techniques, including generative adversarial networks (GANs) trained to produce realistic wound images with controlled variations in tissue composition, size, and imaging conditions, may supplement organic data collection to address specific performance gaps identified during validation studies.

Model architecture refinements will explore advanced deep learning techniques that have demonstrated superior performance in medical imaging domains. Vision transformer architectures, which leverage self-attention mechanisms to capture long-range spatial dependencies, may enhance tissue segmentation accuracy for large, heterogeneous wounds where conventional convolutional approaches struggle with context integration. Multi-task learning frameworks that jointly optimize classification, segmentation, and measurement objectives through shared feature representations could improve parameter efficiency and exploit synergies between related tasks. Additionally, uncertainty quantification mechanisms, such as Monte Carlo dropout, deep ensembles, or evidential deep learning could provide confidence estimates accompanying algorithmic predictions, enabling clinicians to differentiate high-confidence assessments requiring minimal review from uncertain cases warranting scrutiny.

#### Expansion to Specialized Wound Types and New Modalities

While the current version of the platform addresses common chronic wound etiologies, significant opportunities exist for expansion into specialized wound types with distinct assessment requirements and clinical considerations. Burns and frostbites representa particularly compelling targets for AI-augmented assessment given the critical importance of rapid, accurate depth classification (superficial, superficial partial-thickness, deep partial-thickness, full-thickness) in determining appropriate treatment urgency and surgical planning. Integrating multispectral imaging modalities, such as near-infrared spectroscopy or laser Doppler perfusion imaging with deep learning analysis could enhance burn depth discrimination beyond what is achievable with conventional RGB photography alone, potentially improving triage accuracy and reducing unnecessary surgical interventions or delayed excisions.

Similarly, surgical site infections (SSIs) represent a high-impact application domain where early detection through automated surveillance could meaningfully improve outcomes and reduce healthcare costs. Machine learning models trained to detect subtle early signs of infection, erythema extending beyond incision margins, serous drainage, wound edge separation, could facilitate proactive intervention before progression to deep tissue infections requiring reoperation.

#### Global Health Applications and Accessibility

The burden of chronic wounds disproportionately affects underserved populations globally, including individuals in low- and middle-income countries (LMICs), rural communities with limited access to specialized wound care, and economically disadvantaged populations facing structural healthcare barriers. The mobile-first architecture and minimal hardware requirements of the wounds platform position it well for deployment in resource-constrained settings where smartphone penetration increasingly outpaces access to specialized medical equipment or wound care expertise.

Future development will emphasize features specifically designed to enhance accessibility and equity in wound care delivery. Offline functionality, enabling local inference without requiring continuous internet connectivity, is essential for deployment in areas with unreliable network infrastructure. Model compression techniques, including quantization and knowledge distillation, will enable deployment on lower-specification mobile devices without sacrificing accuracy. Multilingual interfaces and voice-guided imaging instructions, potentially leveraging automatic speech recognition and real-time translation, could reduce literacy barriers and expand accessibility to non-English-speaking populations.

Telemedicine integration represents a particularly impactful application in geographically dispersed or medically underserved regions. Community health workers or family caregivers, equipped with the application, could capture standardized wound assessments that are transmitted to remote wound care specialists for asynchronous review and treatment guidance. This store-and-forward telemedicine model, combined with the AI system’s preliminary triage and recommendation capabilities, could extend specialized wound care expertise to populations currently lacking access while optimizing specialist time allocation toward cases requiring nuanced clinical judgment.

Importantly, equitable AI deployment requires proactive assessment and mitigation of potential biases that could exacerbate healthcare disparities. Systematic evaluation of model performance across demographic subgroups, stratified by age, sex, race, ethnicity, skin phototype, and socioeconomic indicators, is essential to identify and address differential accuracy that could disadvantage marginalized populations. Participatory design methodologies, engaging diverse stakeholders including patients, community health workers, and frontline clinicians from varied care settings in the development process, will help ensure that the platform meets the needs and respects the values of the communities it aims to serve.

In conclusion, the wounds platform represents a meaningful step toward transforming wound assessment from a subjective, manual process into a standardized, data-driven discipline. By uniting quantitative imaging, interpretable artificial intelligence, and human clinical expertise within an accessible mobile application, the system addresses longstanding barriers to consistent wound care while respecting the irreplaceable role of clinician judgment. The path forward encompassing prospective validation, EMR integration, treatment personalization, global health applications, and regulatory navigation, is substantial but clearly delineated. As the platform evolves through iterative refinement informed by real-world deployment experience, it holds promise not only to improve individual patient outcomes but to generate the rich, longitudinal datasets necessary to advance wound science itself. The vision of accessible, precision wound care grounded in objective assessment and evidence-based decision support is within reach; realizing this vision requires sustained commitment to rigorous validation, inclusive design, and continuous learning from clinical practice.

## Data Availability

All Data for this study is not available due to patient-consent for scope-of-use.

